# Body Mass Index and breastfeeding behavior among women participants in the Million Veteran Program

**DOI:** 10.1101/2024.07.02.24309047

**Authors:** Joanna Lankester, Rodrigo Guarischi-Sousa, Austin T Hilliard, VA Million Veteran Program, Labiba Shere, Marya Husary, Susan Crowe, Philip S Tsao, David H Rehkopf, Themistocles L Assimes

## Abstract

**Background:** Breastfeeding has established health benefits for infants and has been associated with postpartum improved maternal cardiometabolic health in the long term. However, breastfeeding prevalence is also inversely associated with prepartum body mass index (BMI), and both are linked to socioeconomic factors. We sought to clarify the relationship between prepartum BMI and breastfeeding prevalence in the Million Veteran Program (MVP), a large-scale genetic epidemiology study of US Veterans.

**Methods:** We included data from parous female participants with available breastfeeding information from the MVP cohort. BMI at enrollment as well as earliest BMI available were extracted from the electronic health record, and polygenic scores (PGS) for BMI were calculated for the subset of participants with genotype data. We modeled whether participants breastfed an infant for one month or more (BF≥1M) as a function of BMI at enrollment (n=20,293); earliest BMI where available pre-pregnancy (n=532); and PGS for BMI among genetically inferred European ancestry participants (n=11,568). We conducted Mendelian randomization for breastfeeding using PGS as an instrumental variable.

**Results:** A higher BMI predicted a lower likelihood of BF≥1M in all analyses. A +5 kg/m^2^ BMI pre-pregnancy was associated with a 24% reduced odds of BF≥1M, and a +5 kg/m^2^ genetically predicted BMI was associated with a 17% reduced odds of BF≥1M.

**Conclusions:** BMI predicts a lower likelihood of BF≥1M. Given the high success of breastfeeding initiation in supportive environments combined with potential health benefits to both infant and mother, pregnant Veterans with prepartum elevated BMI may benefit from additional postpartum breastfeeding support.

---

Background

Breastfeeding is associated with maternal health benefits, such as lower incidence of type 2 diabetes, in addition to well established benefits for infants. Body mass may have a complex time-dependent relationship with breastfeeding. Some studies have found that breastfeeding is associated with greater postpartum weight loss, while others have found no effect^1^. In the other direction, body mass prepartum may impact breastfeeding. Prospective studies have shown that those who breastfeed, or breastfeed for a longer duration, already had a lower pre-pregnancy body mass index (BMI)^2, 3^.

Potentially confounding this relationship are multiple socioeconomic factors. Women who breastfeed are more likely to be married and highly educated; they have a higher average household income, and lower rates of smoking and alcohol use^4-6^. Breastfeeding is also associated with race and with both personal and structural experiences with racism^7, 8^. Therefore, breastfeeding may be in part a marker of overall health status and social determinants of health^9^. The association between BMI and breastfeeding could merely reflect practicalities such as work obligations that interfere with breastfeeding.However, this association has held even in a population with substantial postpartum work leave^10^, suggesting the presence of plausible biologic factors such as a delayed onset of lactogenesis that has been observed among overweight and obese mothers could be a factor^11^. Delayed lactogenesis has previously been associated with cessation of breastfeeding by 4 weeks postpartum in the Infant Feeding Practices Study^12^. Lactogenesis II (the onset of copious milk secretion) is driven by a drop in progesterone with placental ejection and occurs regardless of attempting breastfeeding^13^. A delayed onset of lactation may lead to formula supplementation, which in turn may decrease the removal of milk from the breast, interfering with the supply-and-demand mechanism that drives the steady-state lactogenesis III by day 10 postpartum. Thus, delayed lactogenesis may ultimately truncate duration of breastfeeding.

Use of a polygenic score (PGS) can mitigate some of the effects of confounding given that genetic variation is randomly assigned at conception and is less likely to be linked to confounders. A PGS sums the effect size-weighted genotypes of an individual to give an individual’s genetic liability to a health-related trait or disease. Large genetic data sources are required for the development of a PGS, and therefore these scores are typically evaluated on samples that include individuals of a variety of demographic characteristics, such as age, ancestry, and sex^14^. When using PGS in analysis, ancestry is accounted for to avoid population stratification.

In this study, we examine the association between BMI and breastfeeding behavior in a convenience sample of female US Veterans participating in a large-scale genetic epidemiology study who provided historical maternal health information through a baseline questionnaire as well as access to historically measured weights through one of the nation’s oldest integrated electronic health record systems established nearly three decades ago. Our primary aim was to understand how prepartum BMI affects breastfeeding behavior through the use of pre-pregnancy measurements and causal inference methods.

## Methods

### Data source and population

The Million Veteran Program (MVP) is an observational cohort study and biobank of US Veterans enrolled from 2011 to the present^15^. Any Veteran who served in the active military, naval, air, or space service and is eligible for care within the Department of Veterans Affairs (VA) health care system after discharge may enroll in MVP after providing consent for blood banking and linking their genomic data to their health information in the VA electronic health record (EHR). Data for this study were obtained from the optional Baseline and Lifestyle Surveys, the EHR, and genotype data from DNA extracted from blood samples drawn after enrollment. Participants in this study were enrolled between January 2011 and September 2021 and had completed the Lifestyle Survey section on sex-specific female reproductive health questions with non-missing and non-zero values for the number of live births, as well as non-missing values for breastfeeding history.

### Outcome and exposures

Among Veterans reported having given birth one or more times, we used the question in the Lifestyle Survey, “Did you breastfeed your child(ren) for a least 1 month?”, hereafter referred to as breastfed for one or more month (BF≥1M). Historical height, weight, and BMI information was extracted from the EHR close to the time of enrollment and compared to the analogous self-reported data Baseline Survey data. The Baseline Survey value was used if it existed and the values agreed with EHR data, but otherwise the data from the EHR was used^16^. We also extracted directly from the EHR the earliest weight and height measured and the age at this measurement to obtain the earliest BMI. We used Python 3.9.1 for data extraction/transformation and R 4.0.3 for data analyses.

### Covariates

We obtained covariates of age, race, ethnicity, education, income, smoking status, and marital/partner status from the Baseline Survey. We obtained the number of children born, breastfeeding history, and age at first childbirth from the Lifestyle Survey. Details of these covariates are listed in Supplementary File Table S1. Summary statistics were extracted for covariates and breastfeeding history as a function of BMI variables.

### Statistical Analysis

#### Observational associations

We modeled BF≥1M as a function of BMI using logistic regression. BMI was considered both as a continuous variable and by quartile. We used the two measures of BMI as previously described – BMI at enrollment and earliest BMI. We stratified the subset of participants whose earliest BMI was prior to childbirth as a sensitivity analysis due to the potential for reverse causation. We additionally modeled BMI in this subset according to category of earliest BMI (underweight, normal, overweight, obese), though the quartile analysis was the primary analysis to maximize statistical power. Each analysis was conducted with three sets of covariates: (1) as a univariate analysis; (2) adjusted for age (at the date of the respective BMI value), race/ethnicity, education, income, smoking; (3) additionally adjusted for married/partnered status and number of children born.

#### Genetic association and instrumental variable analysis

We used the PGS catalog entry 27 (https://www.pgscatalog.org/score/PGS000027/) to calculate a BMI polygenic score (PGS) in MVP participants. The entry included a set of weights for 2,100,302 genetic variants, or single nucleotide polymorphisms (SNPs) derived from a genome wide association study of 238,944 individuals and validated in an independent data set of 119,951^17^. Both the training and validation datasets that generated the list of SNPs and weights for the PGS were independent of MVP. Genetic data in MVP originated from a custom Axiom array based on the Axiom Genotyping Platform and was imputed using the TOPMed panel in GRCh38/hg38^18^. A PGS for each individual was calculated as the sum of the products of imputed genetic dosage at each SNP and the corresponding PGS catalog weight^19^, with a higher score representing a genetic liability for a higher BMI. Major ancestry groups had previously been determined based on Genetically Inferred Ancestry^20^. We standardized the PGS within each stratified ancestry group.

Population membership was defined based on a genotyped Veteran’s genetically inferred ancestry (GIA) rather than their self-identified race/ethnicity using a reference dataset of unrelated individuals from the 1000 Genomes Project (1KGP)^20^. This assignment was performed centrally as part of a core MVP project and made available as a core resource for all MVP investigators^20^. We standardized the PGS within each stratified ancestry group and verified the presence of an association between the PGS and BMI. Within each GIA stratum, we evaluated the BF≥1M outcome as a function of the PGS, both continuously and by quartile., adjusting for age at enrollment and the first 10 genetic principal components.

We further used the PGS as an instrumental variable for a one-sample Mendelian Randomization analysis. We conducted a Wald ratio analysis within each major ancestry stratum, adjusting for the first 10 genetic principal components, using the R MendelianRandomization package.

## Results

### Summary statistics

The data include 20,375 parous women eligible for inclusion in this study, with BMI at enrollment available for 20,293 and BMI at or prior to their first birth available for 532 (Supplementary File Figure S2). A total of 12,315 (60.7%) of participants had BF≥1M. Among the included Veterans with and without genetic data, higher BMI at enrollment was associated with a younger age of enrollment, self-identified Black or African American race, education less than a Bachelor’s degree, low or medium income level, a report of being a former smoker, and non-response to marital status and smoking survey questions, while it was inversely associated with a report of being a current smoker and being married/partnered (Table 1a).

**Table 1a:** Covariates and outcome by body mass index(BMI) quartile (Q1 - lowest value, Q4 - highest value) and in total. Count (percentage) or mean (standard deviation).

|  |  |  |  |  |  |  |  |
| --- | --- | --- | --- | --- | --- | --- | --- |
| **Variable** | **Value or units** | **Q1** | **Q2** | **Q3** | **Q4** | **Total** | **p value** |
| Number of participants | - | 5074 | 5073 | 5073 | 5073 | 20293 | n/a |
| BMI | kg/m<sup>2</sup> | 22.41 ( 1.93 ) | 27.1 ( 1.13 ) | 31.2 ( 1.33 ) | 38.74 ( 4.64 ) | 29.87 (6.56) | n/a |
| Race/ethnicity | Black | 696 (13.72%) | 1069 (21.07%) | 1193 (23.52%) | 1198 (23.62%) | 4156 (20.48%) | 1.46E-45 |
|  | Hispanic | 348 (6.86%) | 436 (8.59%) | 416 (8.20%) | 348 (6.86%) | 1548 (7.63%) |  |
|  | White | 3803 (74.95%) | 3357 (66.17%) | 3275 (64.56%) | 3348 (66.00%) | 13783 (67.92%) |  |
|  | Other | 227 (4.47%) | 211 (4.16%) | 189 (3.73%) | 179 (3.53%) | 806 (3.97%) |  |
| Education level | No college degree | 1640 (32.32%) | 1627 (32.07%) | 1655 (32.62%) | 1775 (34.99%) | 6697 (33.00%) | 1.92E-48 |
|  | Associates degree | 765 (15.08%) | 814 (16.05%) | 896 (17.66%) | 970 (19.12%) | 3445 (16.98%) |  |
|  | Bachelors degree | 1050 (20.69%) | 1073 (21.15%) | 986 (19.44%) | 896 (17.66%) | 4005 (19.74%) |  |
|  | Post-bachelors degree | 1026 (20.22%) | 872 (17.19%) | 759 (14.96%) | 559 (11.02%) | 3216 (15.85%) |  |
|  | No answer | 593 (11.69%) | 687 (13.54%) | 777 (15.32%) | 873 (17.21%) | 2930 (14.44%) |  |
| Income tertile | Low | 1292 (25.46%) | 1274 (25.11%) | 1337 (26.36%) | 1545 (30.46%) | 5448 (26.85%) | 8.71E-31 |
|  | Middle | 1236 (24.36%) | 1278 (25.19%) | 1344 (26.49%) | 1383 (27.26%) | 5241 (25.83%) |  |
|  | High | 1498 (29.52%) | 1445 (28.48%) | 1260 (24.84%) | 1004 (19.79%) | 5207 (25.66%) |  |
|  | No answer | 1048 (20.65%) | 1076 (21.21%) | 1132 (22.31%) | 1141 (22.49%) | 4397 (21.67%) |  |
| Marital/partner status | Currently partnered or married | 3543 (69.83%) | 3412 (67.26%) | 3350 (66.04%) | 3282 (64.70%) | 13587 (66.95%) | 1.96E-10 |
|  | Formerly married | 780 (15.37%) | 800 (15.77%) | 783 (15.43%) | 793 (15.63%) | 3156 (15.55%) |  |
|  | Never married | 125 (2.46%) | 144 (2.84%) | 143 (2.82%) | 114 (2.25%) | 526 (2.59%) |  |
|  | No answer | 626 (12.34%) | 717 (14.13%) | 797 (15.71%) | 884 (17.43%) | 3024 (14.90%) |  |
| Smoking status | Never | 1994 (39.30%) | 2134 (42.07%) | 2074 (40.88%) | 2061 (40.63%) | 8263 (40.72%) | 4.84E-30 |
|  | Former | 1496 (29.48%) | 1515 (29.86%) | 1536 (30.28%) | 1623 (31.99%) | 6170 (30.40%) |  |
|  | Current | 1038 (20.46%) | 776 (15.30%) | 738 (14.55%) | 559 (11.02%) | 3111 (15.33%) |  |
|  | No answer | 546 (10.76%) | 648 (12.77%) | 725 (14.29%) | 830 (16.36%) | 2749 (13.55%) |  |
| Age | years | 56.09 ( 13.94 ) | 55.97 ( 12.63 ) | 55.75 ( 11.78 ) | 55.5 ( 10.88 ) | 55.83 (12.36) | 0.00963 |
| Number of births | - | 2.16 ( 1.15 ) | 2.21 ( 1.15 ) | 2.19 ( 1.16 ) | 2.2 ( 1.15 ) | 2.19 (1.15) | 0.187 |
| Polygenic score for BMI (scaled within each ancestry group) | - | -0.28 ( 1.0 ) | -.08 ( 0.99 ) | 0.04 ( 0.97 ) | 0.27 ( 0.99 ) | -0.010 (1.01) | 6.51E-142 |
| Breastfed for 1 month or more | No | 1807 (35.61%) | 1906 (37.57%) | 2100 (41.40%) | 2165 (42.68%) | 7978 (39.31%) | 7.53E-15 |
|  | Yes | 3267 (64.39%) | 3167 (62.43%) | 2973 (58.60%) | 2908 (57.32%) | 12315 (60.69%) |  |

### Prediction of BMI by polygenic score

BMI was strongly associated with the within-ancestry scaled PGS for BMI (p=6.51 x 10^-142^) and was inversely associated with having BF≥1M (p=7.53 x 10^-13^). The mean BMI was 29.9 kg/m^2^ (standard deviation 6.6), with means by quartile of 22.4, 27.1, 31.2, and 38.7 (Table 1b). A standard deviation increase in the PGS was responsible for an increase in BMI at enrollment of 0.8-1.5 kg/m^2^ and an increase in earliest pre-pregnancy BMI of 0.8-1.2 kg/m^2^, depending on the ancestry group (Table 1b-Table 1c). The PGS accounted for 1.7-5.1% of variation in BMI at enrollment and 2.4-7.3% of variation in prepartum BMI (Table 1b-Table 1c). The PGS performed better in participants of European ancestry, consistent with the predominantly European ancestry background of the training dataset that had been used to generate the PGS catalog entry.

**Table 1b:** Relationship between polygenic score (PGS) and body mass index(BHI) at enrollment: by quartile (Q1 - lowest value, Q4 - highest value) and continuous. Mean (standard deviation) of BMI (kg/m^2) by PGS quartile. Coefficient, p-value, and R^2 for the increase in BMI corresponding to 1 standard deviation of increase in PGS.

| Group | n | Mean (SD) of BMI (kg/m<sup>2</sup>) by PGS quartile |  |  |  | Effect of continuously measured PGS on BMI |  |  |
| --- | --- | --- | --- | --- | --- | --- | --- | --- |
|  |  | Q1 (lowest PGS) | Q2 | Q3 | Q4 (highest PGS) | Continuous (kg/m<sup>2</sup> increase per SD of PGS) | p value (continuous) | R<sup>2</sup> (continuous) |
| EUR | 11568 | 27.7 (6.1) | 29.1 (6.4) | 30.2 (6.7) | 31.6 (7.3) | 1.53 | 2.72E-134 | 0.051 |
| AFR | 3589 | 29.7 (5.8) | 30.6 (6.2) | 31.2 (6.5) | 31.7 (6.1) | 0.811 | 4.76E-15 | 0.017 |
| AMR | 1327 | 28.0 (5.8) | 29.9 (6.3) | 29.9 (5.5) | 31.2 (6.2) | 1.20 | 3.22E-13 | 0.039 |

**Table 1c:** Relationship between polygenic score (PGS) and earliest pre-partum body mass index(BMI): Coefficient, p-value, and R^2 for the Increase in BHI corresponding to 1 standard deviation of Increase In PGS.

| Group | n | Continuous (kg/m<sup>2</sup> increase per SD of PGS) | p value | R<sup>2</sup> |
| --- | --- | --- | --- | --- |
| EUR | 310 | 1.22 | 7.94E-07 | 0.073 |
| AFR | 63 | 1.00 | 0.0737 | 0.036 |
| AMR | 60 | 0.815 | 0.122 | 0.024 |

### Observational analysis: BMI at enrollment

The observational analysis (n=20,293) revealed that a higher BMI at enrollment was associated with a lower rate of BF≥1M across all covariate models, whether BMI was considered continuously or by quartile (Table 2a, Table S3, Table S4; odds ratio (OR) = 0.93; 95% confidence interval [0.91, 0.95] per +5 kg/m^2^ BMI for the fully adjusted model). Quartile estimates decreased monotonically (Q1, OR=1 (reference); Q2, OR=0.95 [0.86, 1.04]; Q3, 0.81 [0.74, 0.89]; Q4, 0.77 [0.70, 0.84]).

**Table 2a:** Observational analysis predicting breastfed for 1 month or more as a function of body mass index(BMI) at enrollment: by quartile (Q1 - lowest value, Q4 - highest value) and continuous (per+5 kg/m^2). Odds ratio and 95% confidence interval.

| Covariates | n | Q1 | Q2 | Q3 | Q4 | Continuous |
| --- | --- | --- | --- | --- | --- | --- |
| None (univariate) | 20293 | ref. (1) | 0.91 (0.84,0.98) | 0.78 (0.72,0.84) | 0.74 (0.68,0.80) | 0.92 (0.90,0.94) |
| age + race/ethnicity + education level + income level + smoking status | 17544 | ref. (1) | 0.97 (0.88,1.06) | 0.83 (0.75,0.90) | 0.78 (0.71,0.85) | 0.93 (0.91,0.95) |
| age + race/ethnicity + education level + income level + smoking status + married/partnered status + number of births | 17544 | ref. (1) | 0.95 (0.86,1.04) | 0.81 (0.74,0.89) | 0.77 (0.70,0.84) | 0.93 (0.91,0.95) |

### Observational analysis: prepartum BMI

In the subset of participants with a recorded BMI at or before the year of birth of the first offspring (n=532), effect sizes were larger with OR=0.76 [0.58, 1.00] per +5 BMI for the fully adjusted model (Table 2b, Table S5, Table S6). The fourth quartile showed an especially pronounced inverse association with BF≥1M (OR=0.34 [0.13, 0.87]). Results by earliest BMI category were consistent (Table S2).

**Table 2b:** Observational analysis predicting breastfed for 1 month or more as a function of earliest prepartum body mass index (BMI): by quartile (Q1 - lowest value, Q4 - highest value) and continuous (per +5 kg/m^2). Odds ratio and 95% confidence interval.

| Covariates | n | Q1 | Q2 | Q3 | Q4 | Continuous |
| --- | --- | --- | --- | --- | --- | --- |
| None (univariate) | 532 | ref. (1) | 0.91 (0.52,1.58) | 0.63 (0.34,1.17) | 0.37 (0.16,0.84) | 0.72 (0.56,0.92) |
| age at earliest BMI measurement + race/ethnicity + education level + income level + smoking status | 529 | ref. (1) | 0.87 (0.48,1.55) | 0.74 (0.38,1.43) | 0.37(0.15,0.94) | 0.76 (0.58,0.99) |
| age at earliest BMI measurement + race/ethnicity + education level + income level + smoking status + married/partnered status + number of births | 529 | ref. (1) | 0.87 (0.48,1.58) | 0.87 (0.44,1.72) | 0.34 (0.13,0.87) | 0.76 (0.58,1.00) |

### Genetic and instrumental variable analysis using polygenic score

Analyses included n=11568, 3589, and 1327 participants of genetically inferred European (EUR), African (AFR), and Admixed American (AMR) ancestry, respectively, with an insufficient count of East Asian ancestry participants (n=131) to generate stable estimates. A higher PGS (and thus higher BMI) was predictive of a lower chance of BF≥1M (Table 2c; OR=0.94 [0.91, 0.98] EUR; 0.94 [0.87, 1.01] AFR; 0.84 [0.73, 0.98] AMR per standard deviation of PGS). Lastly, our Mendelian randomization using the PDG as a genetic instrumental variable showed a decrease in BF≥1M with increased BMI (Table 3; per +5 kg/m^2^ BMI, OR=0.83 [0.74, 0.93] EUR; 0.69 [0.44, 1.09] AFR; 0.62 [0.41, 0.95] AMR).

**Table 2c:** Genetic analysis predicting breastfed for one month or more as a function of polygenic score (PGS) for body mass index (BMI): by quartile (Q1 - lowest value, Q4 - highest value) and continuous (per +1 standard deviation of PGS). Odds ratio and95% confidence Interval. PC=principal component.

| Ancestry group | n | Q1 (lowest PGS) | Q2 | Q3 | Q4 (highest PGS) | Continuous (per +1 SD PGS) |
| --- | --- | --- | --- | --- | --- | --- |
| EUR | 11568 | ref. (1) | 0.89 (0.80,0.99) | 0.82 (0.74, 0.92) | 0.88 (0.79, 0.98) | 0.94 (0.91, 0.98) |
| AFR | 3589 | ref. (1) | 0.91 (0.75, 1.11) | 0.89 (0.73, 1.09) | 0.85 (0.69, 1.04) | 0.94 (0.87, 1.01) |
| AMR | 1327 | ref. (1) | 0.78 (0.55, 1.10) | 0.90 (0.62, 1.30) | 0.70 (0.47, 1.05) | 0.84 (0.73, 0.98) |
| **Covariates: age + PC1 + PC2 + PC3 + PC4 + PC5 + PC6 + PC7 + PC8 + PC9 + PC10** |  |  |  |  |  |  |

**Table 3:**
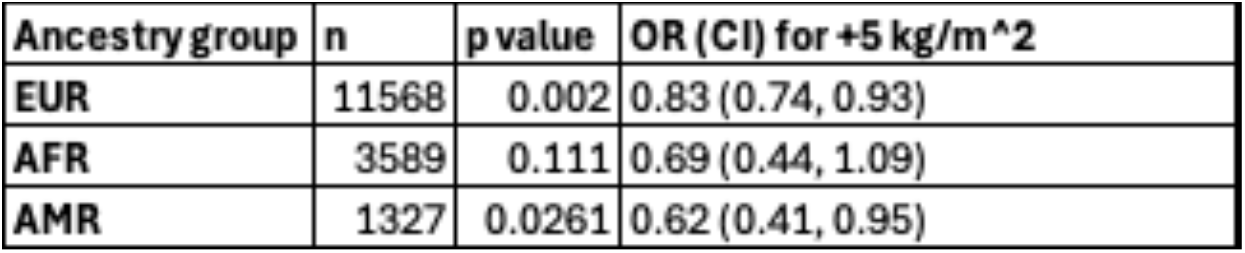
Mendelian randomization predicting breastfed for one month or more as a function of genetically predicted body mass index. P value, odds ratio (OR), and 95% confidence interval(CI) shown.

| Ancestry group | n | p value | OR (CI) for +5 kg/m<sup>2</sup> |
| --- | --- | --- | --- |
| EUR | 11568 | 0.002 | 0.83 (0.74, 0.93) |
| AFR | 3589 | 0.111 | 0.69 (0.44, 1.09) |
| AMR | 1327 | 0.0261 | 0.62 (0.41, 0.95) |

## Discussion

We found that BMI was inversely associated with breastfeeding for our outcome of BF≥1M in all our analyses in this study, mitigating against the potential of reverse causation from a bidirectional relationship between breastfeeding and BMI in two different ways: (1) through the availability of a pre-childbirth measurement of BMI in a subset and (2) via the use of a PGS as an instrumental variable. In the subset of individuals with pre-childbirth BMI available, a +5 kg/m^2^ BMI was associated with a 24% reduced odds of BF≥1M, with a nonlinear trend indicating the highest BMIs led to the greatest risk of not initiating breastfeeding. The Mendelian randomization analysis found the genetic liability for +5 kg/m^2^ BMI associated with at least a 17% reduced odds of BF≥1M.

Our findings suggest the importance of targeted, early breastfeeding support to those with higher BMIs who are at greater risk of being unable to either initiate or sustain breastfeeding. Given that a lack of breastfeeding and a higher BMI have been independently linked with the risk of type 2 diabetes and cardiovascular disease over the long-term^2, 21^, providing such targeted support may not only benefit the infant but also the mother. While our study is restricted to parous Veterans, we have no reason to believe that our findings and the potential benefit cannot be generalized to all parous women.

Our results are consistent with multiple large scale previously published studies examining the relationship between pre-pregnancy body mass and breastfeeding. The MIHA study at UCSF found that pre-pregnancy obesity predicted never breastfeeding^23^. Obesity and overweight status were also associated with lower breastfeeding initiation in the Norwegian MoBa data^24^ and the Australian National Health Survey^25^. A retrospective analysis of over 176,000 women in the North West Thames region showed antenatal overweight/obesity associated with a markedly decreased odds of breastfeeding at discharge^26^. Pre-pregnancy body mass has also been implicated in earlier cessation of breastfeeding. In Nurses Health Study II data, BMI at age 18 years was inversely associated with duration of breastfeeding^2^. Pre-pregnancy BMI was also a predictor of duration of breastfeeding in the Danish National Birth Cohort, despite a nearly universal initiation of breastfeeding^27^.

Plausible biological explanations to explain these associations exist. For example, metabolic profile changes impairing lactogenesis II could be driving the link between decreased breastfeeding and increased BMI. Metabolic factors, including BMI, have been consistently linked to a delay in lactogenesis II. For example, the risk of delayed onset of lactation has been greater with overweight and obese BMI compared to normal range BMI in multiple studies, including a general population from University of California - Davis Medical Center^11^ and a gestational diabetes mellitus specific population in SWIFT^28^. Given BMI is a primary risk factor for dysglycemia, we also note that a study of 26 primiparas at a prenatal clinic found that 56% of the variation in the time to lactogenesis II was explained by a 1-hour post-glucose challenge serum insulin concentration^29^. Finally, a study of 40 women in the rural US found that the serum prolactin response to infant sucking was lower in those within an overweight/obese BMI range than in a normal BMI range^30^. Prolactin is a lactogenic hormone that enables milk production, but it also affects insulin sensitivity and lipid metabolism^31^. Serum prolactin – both during pregnancy^32^ and at 6-9 weeks postpartum^33^ – is inversely associated with later type 2 diabetes. Structural changes in the breast could also be responsible for differences in lactogenesis. For example, a study of diet-induced obese mice found delayed lactogenesis by one day compared to lean mice, with a subset of obese mice from this study showing abnormal alveolar development at day 14 of pregnancy with reduced ductal branching on microscopy of mammary gland tissue cells as compared to non-obese mice^34^.

Researchers have also questioned the direction of effect between breastfeeding and the metabolic syndrome – breastfeeding as a reset of metabolic disruptions from pregnancy vs. metabolic disease adversely affecting breastfeeding such that adverse lactation outcomes are a marker of metabolic dysregulation^9, 35^. We believe both may be true as we have shown that a genetically predicted higher BMI associates with reduced likelihood of BF≥1M. In light of other evidence, we consider that an impaired metabolic profile, as most easily measured by BMI, is responsible for a decrease in breastfeeding.

A strength of our study is the sample size. Although the Veteran population is mostly male, the large number of participants enrolled in MVP resulted in a substantial number of women being eligible for inclusion. Despite the relatively smaller number of participants with prepartum measurements, a sample size of over 500 in this stratum was substantial and sufficient to detect an effect. Nevertheless, replication in additional populations would be warranted. Another strength is diversity, with over 40% of the genetic samples in subjects of non-European ancestry.

A weakness of our study was a limited measure of breastfeeding, with information only on whether breastfeeding occurred for one month or more and no further information on breastfeeding duration. We do not have information on whether breastfeeding was attempted but not sustained vs. never attempted. Moreover, the survey question does not differentiate between multiparas who breastfed all children vs. only one child for one month or more. The survey also did not include information on exclusivity of breastfeeding vs. combined breastfeeding and formula feeding. Further studies are needed in data sets with additional variables. Another weakness was that most measured BMIs occurred after pregnancy, so the observational analysis of BMI at enrollment may suffer from reverse causation.

In conclusion, we have shown that a higher BMI predicts a lower likelihood of breastfeeding for one month or more. Importantly, this finding does not imply that patients with elevated BMI should not attempt breastfeeding; in fact, when adequately supported, breastfeeding initiation can be nearly universal^27^. Instead, our findings suggest additional support should be provided to those with higher BMI in the postpartum period to maximize the probability of successful breastfeeding, which will not only benefit the health of the infant in the short term but also possibly the cardiometabolic health of the mother in the long term.

## Declarations

### Ethics approval and consent to participate

The study received ethical and study protocol approval from the U.S. Veterans Affairs Central Institutional Review Board in accordance with the ethical principles outlined in the Belmont Report and in compliance with the Common Rule (45 CFR 46) and the Final Rule. This study adhered to the Declaration of Helsinki, and informed consent to participate was obtained from all participants.

### Consent for publication

Not applicable

### Data Availability

Access to individual level data used in the present work requires approval by the U.S. Veterans Administration. Genetic summary statistics from Million Veteran Program are available via application through the Database of Genotypes and Phenotypes (dbGaP) accession number phs001672.

### Authors’ contributions

JL conceptualized the study. JL, RG, AH, DHR, and TLA designed the analyses. VAMVP and PST generated the data. JL, RG, AH, LS, and MH wrote software to acquire data subsets or run analyses. JL, RG, AH, SC, DHR, and TLA interpreted the results. JL wrote the initial manuscript draft. JL, MH, SC, DHR, and TLA substantively revised the manuscript. All authors approved the submitted manuscript.

## Acknowledgements

Not applicable

## Consortium members

A full list of consortium details can be found in the Supplement.

Sumitra Muralidhar, Ph.D.^6^, Jennifer Moser, Ph.D.^6^, Jennifer E. Deen, B.S. ^6^, Philip S. Tsao, Ph.D.^7^, Sumitra Muralidhar, Ph.D.^6^, J. Michael Gaziano, M.D., M.P.H.^8^, Elizabeth Hauser, Ph.D. ^9^, Amy Kilbourne, Ph.D., M.P.H. ^10^, Michael Matheny, M.D., M.S., M.P.H. ^11^, Dave Oslin, M.D. ^12^, Deepak Voora, M.D.^9^, Jessica V. Brewer, M.P.H. ^8^, Mary T. Brophy M.D., M.P.H. ^8^, Kelly Cho, M.P.H, Ph.D. ^8^, Lori Churby, B.S. 7, Scott L. DuVall, Ph.D. ^13^, Saiju Pyarajan Ph.D. ^8^, Robert Ringer, Pharm.D. ^14, Luis E. Selva, Ph.D. 8^, Shahpoor (Alex) Shayan, M.S. ^8^, Brady Stephens, M.S. ^15^, Stacey B. Whitbourne, Ph.D. ^8^, Themistocles L. Assimes, M.D., Ph.D. ^7^, Adriana Hung, M.D. ^11^, M.P.H., Henry Kranzler, M.D. ^12^

6. US Department of Veterans Affairs, 810 Vermont Avenue NW, Washington, DC 20420

7. VA Palo Alto Health Care System, 3801 Miranda Avenue, Palo Alto, CA 94304

8. VA Boston Healthcare System, 150 S. Huntington Avenue, Boston, MA 02130

9. Durham VA Medical Center, 508 Fulton Street, Durham, NC 27705

10. VA HSR&D, 2215 Fuller Road, Ann Arbor, MI 48105

11. VA Tennessee Valley Healthcare System, 1310 24th Ave. South, Nashville, TN 37212

12. Philadelphia VA Medical Center, 3900 Woodland Avenue, Philadelphia, PA 19104

13. VA Salt Lake City Health Care System, 500 Foothill Drive, Salt Lake City, UT 84148

14. New Mexico VA Health Care System, 1501 San Pedro Drive SE, Albuquerque, NM 87108

15. Canandaigua VA Medical Center, 400 Fort Hill Avenue, Canandaigua, NY 14424

