## Supplementary material for "Body Mass Index and breastfeeding behavior among women participants in the Million Veteran Program": Table S1

### Table S1: Phenotype definitions

| Variable | Variable Type | Source | Question Number | Details |
|---|---|---|---|---|
| Breastfeeding for one month or more | Outcome | Lifestyle Survey | 52h | Binary variable |
| Body mass index (BMI) | Exposure | CoreVitals table (EHR and Baseline Survey combined) | n/a | Weight and height in this table had been produced by harmonizing the EHR and Baseline Survey data, giving priority to the Baseline Survey value if the values agreed or if no measured value existed but replacing with the EHR data if the values were contradictory. |
| Age at enrollment | Covariate | Baseline Survey | 1-2 | n/a |
| Earliest BMI | Exposure (for a subset) | EHR | n/a | Weight was extracted as the weight recorded on the earliest day for each participant. In the case of more than one weight measurement on the earliest day, we used the median. Height was not collected at every instance where weight was measured. Since all data was from adulthood, we chose the median height across all measurements. |
| Age at earliest BMI | Covariate | EHR | | Age was extracted according to date of the earliest weight measurement. |
| Race/ethnicity | Covariate | Baseline Survey | 4-5 | Self reported race/ethnicity was combined into a single variable. Value was "Hispanic or Latino" if ethnicity question was answered as such, regardless of race. Otherwise, participants were grouped into "Black or African American", "White", or "Other". (Self reported variable was used only for observational analysis; genetic analyses used genetically inferred ancestry.) |
| Education | Covariate | Baseline Survey | 7 | Education was combined into 4 categories: 1. No college degree, 2. Associate's Degree, 3. Bachelor's Degree, 4. Any degree beyond Bachelor's. |
| Income | Covariate | Baseline Survey | 10 | Income was combined from 10 categories to approximate tertiles. Values in USD/year were: Low - < 30,000; Medium - 30,000 to < 60,000; High - 60,000 or more. |
| Smoking status | Covariate | CoreVitals table (combined from Baseline and Lifestyle Surveys) | 33; 13-14 | Categories were: Never smoker, Former smoker, Current smoker. |
| Marital/Partnered status | Covariate | Baseline Survey | 8 | Partner status was grouped into: Any partnered (Married, civil commitment, cohabiting); Any formerly partnered (Separated, Divoced, Widowed); Not married. |
| Number of children born | Covariate | Lifestyle Survey | 52d | n/a |
| Age at first & last birth | Covariate | Lifestyle Survey | 52f | n/a |

### Figure S2: Sample size inclusions/exclusions

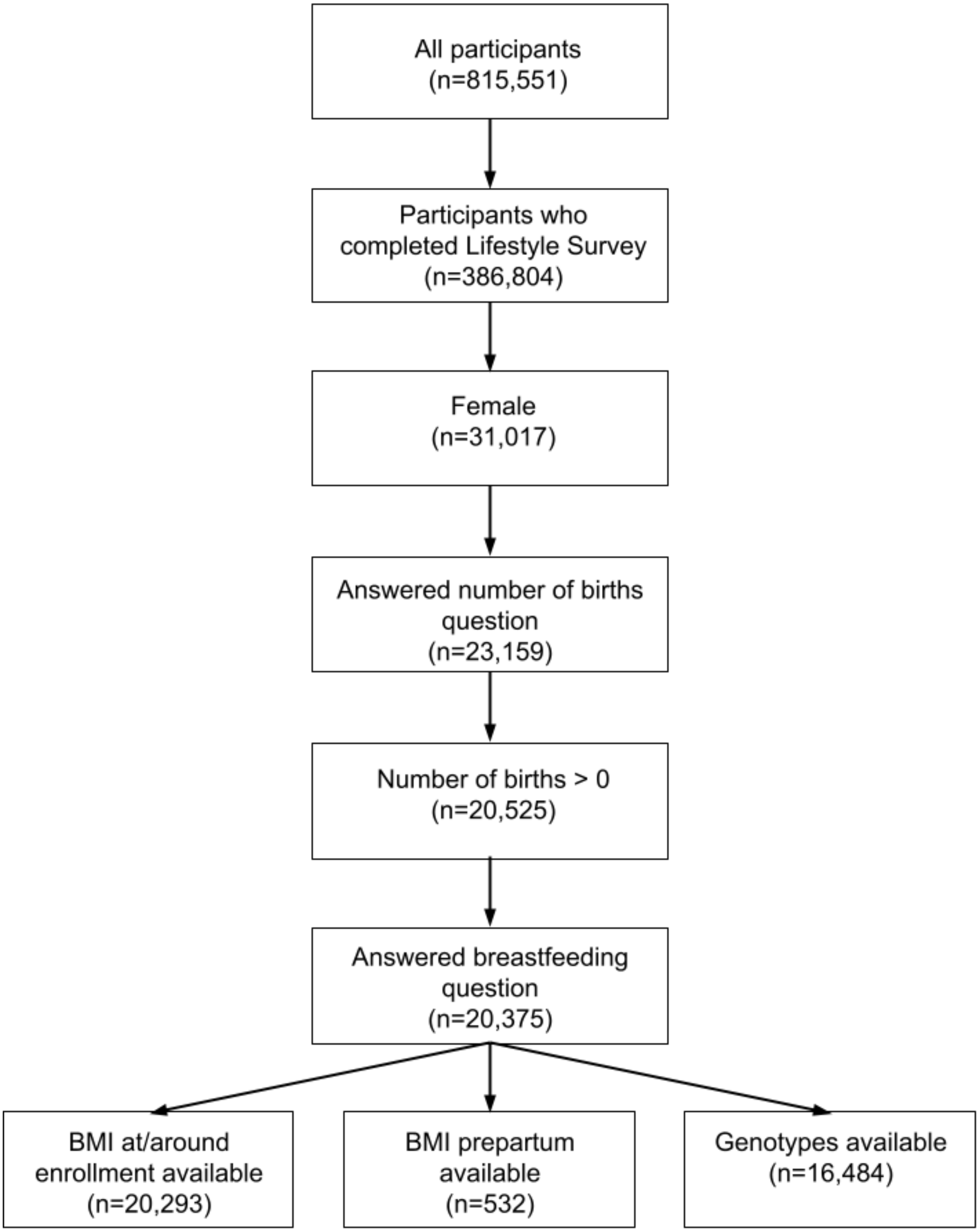

### Table S2: Observational analysis results by category of earliest BMI measurement for participants with pre-pregnancy BMI available

**Table S2: Observational analysis predicting breastfeeding for one month or more as a function of earliest prepartum body mass index (BMI): by category of earliest BMI (Normal is 18.5 to <25 kg/m^2; Overweight is 25 to <30 kg/m^2; Obese is >=30 kg/m^2). Underweight participants (n=7) are omitted as sample size was too small for stable estimates. Odds ratio and 95% confidence interval.**

| Covariates | Normal (n=265) | Overweight (n=185) | Obese (n=75) |
|---|---|---|---|
| None (univariate) | ref. (1) | 0.88 (0.53,1.47) | 0.46 (0.25, 0.84) |
| age at earliest BMI measurement + race/ethnicity + education level + income level + smoking status | ref. (1) | 0.87 (0.50,1.50) | 0.56 (0.29, 1.08) |
| age at earliest BMI measurement + race/ethnicity + education level + income level + smoking status + married/partnered status + number of births | ref. (1) | 0.91 (0.52,1.57) | 0.58 (0.30, 1.14) |

### Table S3: Observational analysis results for all covariates in the model of BMI at enrollment by quartile vs breastfed for one month

<table>
<tr><th colspan="5">Coefficients for all covariates in quartile model of BMI at enrollment vs breastfed 1 month or more</th></tr>
<tr><th></th><th>Estimate</th><th>Std.Error</th><th>z value</th><th>Pr(>|z|)</th></tr>
<tr><td>(Intercept)</td><td>1.030959</td><td>0.111495</td><td>9.247</td><td>< 2e-16 ***</td></tr>
<tr><td>as.factor(bmi_quartile)2</td><td>-0.054754</td><td>0.046299</td><td>-1.183</td><td>0.23696</td></tr>
<tr><td>as.factor(bmi_quartile)3</td><td>-0.209123</td><td>0.046191</td><td>-4.527</td><td>5.97e-06 ***</td></tr>
<tr><td>as.factor(bmi_quartile)4</td><td>-0.260453</td><td>0.046683</td><td>-5.579</td><td>2.42e-08 ***</td></tr>
<tr><td>age</td><td>-0.035169</td><td>0.001505</td><td>-23.363</td><td>< 2e-16 ***</td></tr>
<tr><td>as.factor(raceeth)hispanic</td><td>0.647668</td><td>0.070401</td><td>9.2</td><td>< 2e-16 ***</td></tr>
<tr><td>as.factor(raceeth)other</td><td>1.029847</td><td>0.101752</td><td>10.121</td><td>< 2e-16 ***</td></tr>
<tr><td>as.factor(raceeth)white</td><td>0.673352</td><td>0.042353</td><td>15.899</td><td>< 2e-16 ***</td></tr>
<tr><td>as.factor(education_level)2_medlow</td><td>0.266507</td><td>0.044468</td><td>5.993</td><td>2.06e-09 ***</td></tr>
<tr><td>as.factor(education_level)3_medhigh</td><td>0.513044</td><td>0.044484</td><td>11.533</td><td>< 2e-16 ***</td></tr>
<tr><td>as.factor(education_level)4_highest</td><td>0.844969</td><td>0.0514</td><td>16.439</td><td>< 2e-16 ***</td></tr>
<tr><td>as.factor(education_level)nan</td><td>0.262584</td><td>0.159604</td><td>1.645</td><td>0.09992 .</td></tr>
<tr><td>as.factor(income_level)2_med</td><td>-0.031892</td><td>0.0418</td><td>-0.763</td><td>0.44549</td></tr>
<tr><td>as.factor(income_level)3_high</td><td>0.053364</td><td>0.046269</td><td>1.153</td><td>0.24878</td></tr>
<tr><td>as.factor(income_level)nan</td><td>-0.049611</td><td>0.060752</td><td>-0.817</td><td>0.41415</td></tr>
<tr><td>as.factor(smoker_survey)2</td><td>-0.079438</td><td>0.036863</td><td>-2.155</td><td>0.03116 *</td></tr>
<tr><td>as.factor(smoker_survey)3</td><td>-0.341811</td><td>0.046178</td><td>-7.402</td><td>1.34e-13 ***</td></tr>
<tr><td>as.factor(marital_status)never_married</td><td>0.001773</td><td>0.102474</td><td>0.017</td><td>0.9862</td></tr>
<tr><td>as.factor(marital_status)no_marital_status</td><td>0.166524</td><td>0.141517</td><td>1.177</td><td>0.23931</td></tr>
<tr><td>as.factor(marital_status)partnered</td><td>0.122375</td><td>0.043107</td><td>2.839</td><td>0.00453 **</td></tr>
<tr><td>fbrthqty</td><td>0.302142</td><td>0.015571</td><td>19.404</td><td>< 2e-16 ***</td></tr>
<tr><td></td><td></td><td></td><td></td><td></td></tr>
<tr><td colspan="5">Reference: lowest BMI quartile, self-identified Black race, lowest education level, lowest income level, never smoker, formerly married</td></tr>
<tr><td colspan="5">Signif. codes: 0 ‘***’ 0.001 ‘**’ 0.01 ‘*’ 0.05 ‘.’ 0.1 ‘ ’ 1</td></tr>
</table>

Table S4: Observational analysis results for all covariates in the model of continuous BMI at enrollment vs breastfed for one month

| Coefficients for all covariates in continuous model of BMI at enrollment vs breastfed 1 month or more | | | | | |
|---|---|---|---|---|---|
| | **Estimate** | **Std.Error** | **z value** | **Pr(>\|z\|)** | |
| (Intercept) | 1.754579 | 0.126461 | 13.874 | < 2e-16 *** | |
| bmi | -0.014139 | 0.002481 | -5.698 | 1.21E-08 | *** |
| age | -0.028918 | 0.00142 | -20.361 | < 2e-16 *** | |
| as.factor(raceeth)hispanic | 0.687724 | 0.069442 | 9.904 | < 2e-16 *** | |
| as.factor(raceeth)other | 1.065357 | 0.100471 | 10.604 | < 2e-16 *** | |
| as.factor(raceeth)white | 0.694453 | 0.041391 | 16.778 | < 2e-16 *** | |
| as.factor(education_level)2_medlow | 0.244455 | 0.043906 | 5.568 | 2.58E-08 | *** |
| as.factor(education_level)3_medhigh | 0.464567 | 0.043847 | 10.595 | < 2e-16 *** | |
| as.factor(education_level)4_highest | 0.766945 | 0.050556 | 15.17 | < 2e-16 *** | |
| as.factor(education_level)nan | 0.287565 | 0.143297 | 2.007 | 0.04477 * | |
| as.factor(income_level)2_med | -0.055603 | 0.041126 | -1.352 | 0.17636 | |
| as.factor(income_level)3_high | 0.037983 | 0.045193 | 0.84 | 0.40065 | |
| as.factor(income_level)nan | -0.086883 | 0.059715 | -1.455 | 0.14568 | |
| as.factor(smoker_survey)2 | -0.098115 | 0.036396 | -2.696 | 0.00702 ** | |
| as.factor(smoker_survey)3 | -0.375034 | 0.045566 | -8.23 | < 2e-16 *** | |
| Reference: self-identified Black race, lowest education level, lowest income level, never smoker, formerly married | | | | | |
| Signif. codes: 0 ‘***’ 0.001 ‘**’ 0.01 ‘*’ 0.05 ‘.’ 0.1 ‘ ’ 1 | | | | | |

Table S5: Observational analysis results for all covariates in the model of prepartum BMI by quartile vs breastfed for one month

Coefficients for all covariates in quartile model of pre-partum BMI vs breastfed 1 month or more

| | Estimate | Std.Error | z value | Pr(>\|z\|) |
|---|---|---|---|---|
| (Intercept) | 0.001452 | 0.850226 | 0.002 | 0.9986 |
| as.factor(earliest_bmi_quartile)2 | -0.140035 | 0.30342 | -0.462 | 0.6444 |
| as.factor(earliest_bmi_quartile)3 | -0.141136 | 0.34841 | -0.405 | 0.6854 |
| as.factor(earliest_bmi_quartile)4 | -1.072465 | 0.477973 | -2.244 | 0.0248 * |
| age_at_earliest_bmi | 0.003016 | 0.025473 | 0.118 | 0.9058 |
| as.factor(raceeth)hispanic | 1.27665 | 0.515428 | 2.477 | 0.0133 * |
| as.factor(raceeth)other | 16.62733 | 868.600394 | 0.019 | 0.9847 |
| as.factor(raceeth)white | 0.501759 | 0.338095 | 1.484 | 0.1378 |
| as.factor(education_level)2_medlow | -0.132706 | 0.365824 | -0.363 | 0.7168 |
| as.factor(education_level)3_medhigh | 0.841805 | 0.356562 | 2.361 | 0.0182 * |
| as.factor(education_level)4_highest | 0.357283 | 0.36916 | 0.968 | 0.3331 |
| as.factor(education_level)nan | 1.161698 | 1.4801 | 0.785 | 0.4325 |
| as.factor(income_level)2_med | -0.944804 | 0.379787 | -2.488 | 0.0129 * |
| as.factor(income_level)3_high | -0.12528 | 0.410935 | -0.305 | 0.7605 |
| as.factor(income_level)nan | -1.071511 | 0.568219 | -1.886 | 0.0593 . |
| as.factor(smoker_survey)2 | 0.715736 | 0.325058 | 2.202 | 0.0277 * |
| as.factor(smoker_survey)3 | -0.002231 | 0.359486 | -0.006 | 0.995 |
| as.factor(marital_status)never_married | -0.708693 | 0.562033 | -1.261 | 0.2073 |
| as.factor(marital_status)no_marital_status | -0.21669 | 0.785894 | -0.276 | 0.7828 |
| as.factor(marital_status)partnered | 0.564169 | 0.342352 | 1.648 | 0.0994 . |
| fbrthqty | 0.447961 | 0.214068 | 2.093 | 0.0364 * |

Reference: lowest BMI quartile, self-identified Black race, lowest education level, lowest income level, never smoker, formerly married

Signif. codes: 0 '***' 0.001 '**' 0.01 '*' 0.05 '.' 0.1 ' ' 1

Table S6: Observational analysis results for all covariates in the model of prepartum continuous BMI vs breastfed for one month

Coefficients for all covariates in continuous model of pre-partum BMI vs breastfed 1 month or more

| | Estimate | Std.Error | z value | Pr(>\|z\|) |
|---|---|---|---|---|
| (Intercept) | 1.354658 | 1.057468 | 1.281 | 0.2002 |
| earliest_bmi | -0.053976 | 0.027833 | -1.939 | 0.0525 . |
| age_at_earliest_bmi | 0.001459 | 0.025361 | 0.058 | 0.9541 |
| as.factor(raceeth)hispanic | 1.305841 | 0.513271 | 2.544 | 0.011 * |
| as.factor(raceeth)other | 16.547372 | 875.384057 | 0.019 | 0.9849 |
| as.factor(raceeth)white | 0.531585 | 0.33434 | 1.59 | 0.1118 |
| as.factor(education_level)2_medlow | -0.13334 | 0.365166 | -0.365 | 0.715 |
| as.factor(education_level)3_medhigh | 0.805502 | 0.355279 | 2.267 | 0.0234 * |
| as.factor(education_level)4_highest | 0.340838 | 0.3678 | 0.927 | 0.3541 |
| as.factor(education_level)nan | 1.094806 | 1.48352 | 0.738 | 0.4605 |
| as.factor(income_level)2_med | -0.948652 | 0.378402 | -2.507 | 0.0122 * |
| as.factor(income_level)3_high | -0.121524 | 0.409398 | -0.297 | 0.7666 |
| as.factor(income_level)nan | -1.04819 | 0.566422 | -1.851 | 0.0642 . |
| as.factor(smoker_survey)2 | 0.687345 | 0.322272 | 2.133 | 0.0329 * |
| as.factor(smoker_survey)3 | -0.007772 | 0.358275 | -0.022 | 0.9827 |
| as.factor(marital_status)never_married | -0.698286 | 0.560579 | -1.246 | 0.2129 |
| as.factor(marital_status)no_marital_status | -0.172145 | 0.793403 | -0.217 | 0.8282 |
| as.factor(marital_status)partnered | 0.54121 | 0.340395 | 1.59 | 0.1118 |
| fbrthqty | 0.420205 | 0.212382 | 1.979 | 0.0479 * |

Reference: self-identified Black race, lowest education level, lowest income level, never smoker, formerly married

Signif. codes: 0 '***' 0.001 '**' 0.01 '*' 0.05 '.' 0.1 ' ' 1

### MVP Acknowledgements

**VA Million Veteran Program:**
**Core Acknowledgements for Publications**
**May 2024**

**MVP Program Office**

- Sumitra Muralidhar, Ph.D., Program Director
  US Department of Veterans Affairs, 810 Vermont Avenue NW, Washington, DC 20420
- Jennifer Moser, Ph.D., Associate Director, Scientific Programs
  US Department of Veterans Affairs, 810 Vermont Avenue NW, Washington, DC 20420
- Jennifer E. Deen, B.S., Associate Director, Cohort & Public Relations
  US Department of Veterans Affairs, 810 Vermont Avenue NW, Washington, DC 20420

**MVP Executive Committee**

- Co-Chair: Philip S. Tsao, Ph.D.
  VA Palo Alto Health Care System, 3801 Miranda Avenue, Palo Alto, CA 94304
- Co-Chair: Sumitra Muralidhar, Ph.D.
  US Department of Veterans Affairs, 810 Vermont Avenue NW, Washington, DC 20420
- J. Michael Gaziano, M.D., M.P.H.
  VA Boston Healthcare System, 150 S. Huntington Avenue, Boston, MA 02130
- Elizabeth Hauser, Ph.D.
  Durham VA Medical Center, 508 Fulton Street, Durham, NC 27705
- Amy Kilbourne, Ph.D., M.P.H.
  VA HSR&D, 2215 Fuller Road, Ann Arbor, MI 48105
- Michael Matheny, M.D., M.S., M.P.H.
  VA Tennessee Valley Healthcare System, 1310 24th Ave. South, Nashville, TN 37212
- Dave Oslin, M.D.
  Philadelphia VA Medical Center, 3900 Woodland Avenue, Philadelphia, PA 19104
- Deepak Voora, MD
  Durham VA Medical Center, 508 Fulton Street, Durham, NC 27705

**MVP Co-Principal Investigators**

- J. Michael Gaziano, M.D., M.P.H.
  VA Boston Healthcare System, 150 S. Huntington Avenue, Boston, MA 02130
- Philip S. Tsao, Ph.D.
  VA Palo Alto Health Care System, 3801 Miranda Avenue, Palo Alto, CA 94304

**MVP Core Operations**

- Jessica V. Brewer, M.P.H., Director, MVP Cohort Operations
  VA Boston Healthcare System, 150 S. Huntington Avenue, Boston, MA 02130

- Mary T. Brophy M.D., M.P.H., Director, VA Central Biorepository
  VA Boston Healthcare System, 150 S. Huntington Avenue, Boston, MA 02130
- Kelly Cho, M.P.H, Ph.D., Director, MVP Phenomics
  VA Boston Healthcare System, 150 S. Huntington Avenue, Boston, MA 02130
- Lori Churby, B.S., Director, MVP Regulatory Affairs
  VA Palo Alto Health Care System, 3801 Miranda Avenue, Palo Alto, CA 94304
- Scott L. DuVall, Ph.D., Director, VA Informatics and Computing Infrastructure (VINCI)
  VA Salt Lake City Health Care System, 500 Foothill Drive, Salt Lake City, UT 84148
- Saiju Pyarajan Ph.D., Director, Data and Computational Sciences
  VA Boston Healthcare System, 150 S. Huntington Avenue, Boston, MA 02130
- Robert Ringer, Pharm.D., Director, VA Albuquerque Central Biorepository
  New Mexico VA Health Care System, 1501 San Pedro Drive SE, Albuquerque, NM 87108
- Luis E. Selva, Ph.D., Director, MVP Biorepository Coordination
  VA Boston Healthcare System, 150 S. Huntington Avenue, Boston, MA 02130
- Shahpoor (Alex) Shayan, M.S., Director, MVP PRE Informatics
  VA Boston Healthcare System, 150 S. Huntington Avenue, Boston, MA 02130
- Brady Stephens, M.S., Principal Investigator, MVP Information Center
  Canandaigua VA Medical Center, 400 Fort Hill Avenue, Canandaigua, NY 14424
- Stacey B. Whitbourne, Ph.D., Director, MVP Cohort Development and Management
  VA Boston Healthcare System, 150 S. Huntington Avenue, Boston, MA 02130

**MVP Publications and Presentations Committee**

- Co-Chair: Themistocles L. Assimes, M.D., Ph. D
  VA Palo Alto Health Care System, 3801 Miranda Avenue, Palo Alto, CA 94304
- Co-Chair: Adriana Hung, M.D.; M.P.H
  VA Tennessee Valley Healthcare System, 1310 24th Ave. South, Nashville, TN 37212
- Co-Chair: Henry Kranzler, M.D.
  Philadelphia VA Medical Center, 3900 Woodland Avenue, Philadelphia, PA 19104

### Data from the Veterans Healthcare Administration System of Records

US Department of Veterans Affairs. System of Records Notice 97VA10P1: Consolidated Data Information System-VA. 76 FR 25409. May 4, 2011. Amended March 3, 2015.
